# An evaluation of toxigenic Clostridioides difficile positivity as a patient outcome metric of antimicrobial stewardship in Saudi Arabia

**DOI:** 10.1101/2021.02.23.21252226

**Authors:** Christopher A. Okeahialam, Ali A. Rabaan, Albert Bolhuis

## Abstract

**Background:** Antimicrobial stewardship has been associated with a reduction in the incidence of health care associated Clostridium difficile infection (HA-CDI). However, CDI remains under-recognized in many low and middle-income countries where clinical and surveillance resources required to identify HA-CDI are often lacking. The rate of toxigenic C. difficile stool positivity in the stool of hospitalized patients may offer an alternative metric for these settings, but its utlity remains largely untested.

**Aim/Objective:** To examine the impact of an antimicrobial stewardship on the rate of toxigenic C. difficile positivity among hospitalized patients presenting with diarrhoea

**Methods:** A 12-year retrospective review of laboratory data was conducted to compare the rates of toxigenic C. difficile in diarrhoea stool of patients in a hospital in Saudi Arabia, before and after implementation of an antimicrobial stewardship program

**Result:** There was a significant decline in the rate of toxigenic C difficile positivity from 9.8 to 7.4% following the implementation of the antimicrobial stewardship program, and a reversal of a rising trend.

**Discussion:** The rate of toxigenic C. difficile positivity may be a useful patient outcome metric for evaluating the long term impact of antimicrobial stewardship on CDI, especially in settings with limited surveillance resources. The accuracy of this metric is however dependent on the avoidance of arbitrary repeated testing of a patient for cure, and testing only unformed or diarrhoea stool specimens. Further studies are required within and beyond Saudi Arabia to examine the utility of this metric.

## Background

*Clostridioides difficile* infection (CDI) is caused by toxigenic strains of C. *difficile* (tCd), a Gram-positive, spore forming, antibiotic-resistant anaerobic bacillus. CDI is considered to be the most common cause of diarrhoea in healthcare settings in Europe and North America, and an urgent antimicrobial resistance threat (CDC, 2019).

Most of what is known of the impact of antimicrobial stewardship on CDI has emerged from studies in North America and Europe. Systematic reviews indicate that the incidence of healthcare-associated CDI (HA-CDI) is the metric most often utilized in these studies (Feazel *et al* 2014, Baur *et al* 2017).

Other studies (Talpaert et al 2011, Jump et al 2012, Lawes et al 2017) have measured the rate of tCd positivity in diarrhoea stool as a proxy for CDI, and observed a reduction following the implementation of antimicrobial stewardship interventions. This is a simpler metric than the incidence of HA-CDI because it does not require clinical diagnosis of CDI, and does not distinguish between hospital and community-acquired CDI. It may therefore find broader applicability in settings lacking the level of surveillance and diagnostic capacity required to identify HA-CDI. A potential drawback however is that the rate of tCd positivity in a hospital setting may be inflated by a high level of asymptomatic tCd carriers in the catchment population, thereby obscuring the true impact of hospital-based stewardship interventions.

In Saudi Arabia, the few available CDI studies (Al-Tawfiq and Abed 2010, Alzaharani and Al Johani 2013, Hudhaiah and Elhadi 2019) suggest that the prevalence in Saudi communities may be under-estimated, due to a combination of low diagnostic suspicion and resources. This assertion is supported by Saudi studies on self-medication (non-prescription use) with antibiotics (Al-Mohamadi et al 2013, Alshammari et al 2017, Bin Nafisah *et al* 2017, Alhomoud *et al* 2018) that highlight unregulated and inappropriate population exposure to antibiotics, an important risk factor for tCd/CDI.

The true burden of tCd in Saudi communities remains unknown however, and the extent to which this limits the utility of tCd positivity as a metric of hospital antimicrobial stewardship is uncertain. Given these uncertainties, we decided to compare the period positivity rate of tCd in diarrhoea stool obtained from in-patients before and after the implementation of a hospital antimicrobial stewardship program in Saudi Arabia.

## METHODS

### Setting

Johns Hopkins Aramco Healthcare (JHAH) is a 5-site healthcare facility in the Eastern province of Saudi Arabia. The study setting, Dhahran health centre (DHC) is the largest hospital of JHAH. Services include general medicine and surgery, intensive care, and management of haematological and solid organ malignancies. The hospital serves a catchment population of approximately 370,000 patients.

### Study Design

This was retrospective study covering a twelve-year period. The study compared the rate of tCd positivity among in-patients at JHAH, in the six year intervals before and after implementation of the antimicrobial stewardship program (ASP). The study was approved by the Institutional Research Board of JHAH.

The ASP was formally launched in 2011, starting with education sessions specifically targeted at prescribers and hospital pharmacists (Table 1). This was preceded in 2010 however, by the introduction of restricted laboratory reporting of antibiotic susceptibilities for selected Gram negative organisms (Al-Tawfiq et al, 2015). Other program components were introduced in 2012. These were automatic renal dosing, an antibiotic de-escalation protocol, intravenous to oral conversion program, vancomycin pharmacokinetic monitoring, pre-operative antibiotic protocol/adapted orders and multiple interventions including prospective audit and feedback paediatric outpatient clinics to improve prescribing practices for upper respiratory tract infections in paediatric outpatient clinics (Al-Tawfiq and Alawami 2017). The antimicrobial stewardship program was led by an infection disease physician and pharmacist.

**Table 1.** JHAH Antimicrobial Stewardship Interventions

| Intervention | Date of Implementation |
| --- | --- |
| Prescriber and Pharmacist Education sessions | 2011 |
| Automatic Renal Dosing | 2012 |
| Antibiotic De-escalation protocol | 2012 |
| Intravenous to Oral Conversion Program; | 2012 |
| Vancomycin Pharmacokinetic monitoring | 2012 |
| Peri-operative antibiotic protocols using adapted orders | 2012 |
| Multiple interventions including prospective audits, and feedback to prescribers in paediatric outpatient clinics to improve the use of antibiotics for upper respiratory tract infections | 2012 |

### Outcome

The rates of tCd positivity before and after the implementation of the ASP were estimated by dividing the number of tCd positive patients by the total number of patients tested in each period. Duplicate positive tests were excluded for each patient, except when a second positive test was obtained more than 8 weeks after the first, in which case it was considered a new infection and included in the positivity estimate.

The age group categories used were elderly (65 years and above), adult (18 to 64 years) and paediatric (3-17 years). Paediatric patients below the age of 3 were excluded from the study due to the propensity of the 0-2 age group for asymptomatic colonization with toxigenic and non-toxigenic C. *difficile* strains (American Academy of Paediatrics, 2013, Antonara and Leber 2016)

### Microbiology

The C. difficile policy in the study setting is to test only loose or watery stool (stool conforming to the shape of the container). Criteria for sending diarrhea or loose stool specimens to the laboratory for C.difficile testing are: 3 or more loose/watery bowel movements in a 24 hour period, bowel movements unusual or different for the patient, no other recognized aetiology for the diarrhea (e.g. laxative use, inflammatory bowel disease).

Over the study period, stool specimens were tested for *C. difficile* toxins A and B using Enzyme-linked immunosorbent assay (Premier Toxins A&B EIA; Meridian Bioscience). Test results and anonymised demographic data (patients’ age, gender, and in-patient location at the time of specimen collection) were received from the laboratory.

### Definitions

A case of CDI was defined as a patient with a positive tCd test. Recurrent CDI (relapse) was defined as patient positive for tCd 2-8 weeks after the initial positive test.

Re-infection was defined as patient positive for tCd more than 8 weeks after the initial positive test (CDC/McDonald et al 2007, Cohen et al 2010).

Age-group-specific tCd positivity was obtained by expressing the number of tCd positive patients in a specific age group as a proportion of the number of patients tested in that group.

### Statistical Analysis

Data was analysed using SPSS version 23 (SPSS Inc., Chicago, IL) for Windows. The Fisher exact test was used to test for differences in proportions. The Student t-test was used to compare the means of continuous variables. Two-sided P values less than 0.05 were considered statistically significant.

## RESULTS

Over the study period (2005 to 2016), a total of 3086 inpatients at Johns Hopkins Aramco hospital (JHAH) were tested for tCd. 260 patients tested positive for tCd, giving an overall period positivity rate of 8.4%.

The mean age of the study population and tCd positive patients was 55 and 59 respectively. The majority (143/260; 55%) of tCd positive patients were elderly (65 years and over), although this age group comprised just over 40% of patients tested for tCd over the study period

A total of 12/260 patient were identified with a repeat tCd positive test more than 8 weeks after the first positive test. In keeping with the study case definition (CDC/McDonald *et al* 2007), these were considered new cases of CDI (re-infections) and included in the period positivity estimate. The majority of re-infections (8/12; 66%) were observed among the elderly. A total of 28/260 patients were identified with tCd 2-8 weeks after the initial positive test. These were considered relapses rather than new cases of CDI and excluded from the positivity estimate. The majority (18/28; 64%) of relapses were observed in the elderly.

The upward trend in tCd positivity observed in the six-year period prior to the implementation of antimicrobial stewardship was reversed in the six-year period following the implementation of the program (figure 1). Overall, there was a significant decline (p=0.022) in tCd positivity from 9.8% (129/1318) to 7.4% (131/1768) pre and post ASP respectively.

**Figure 1.**
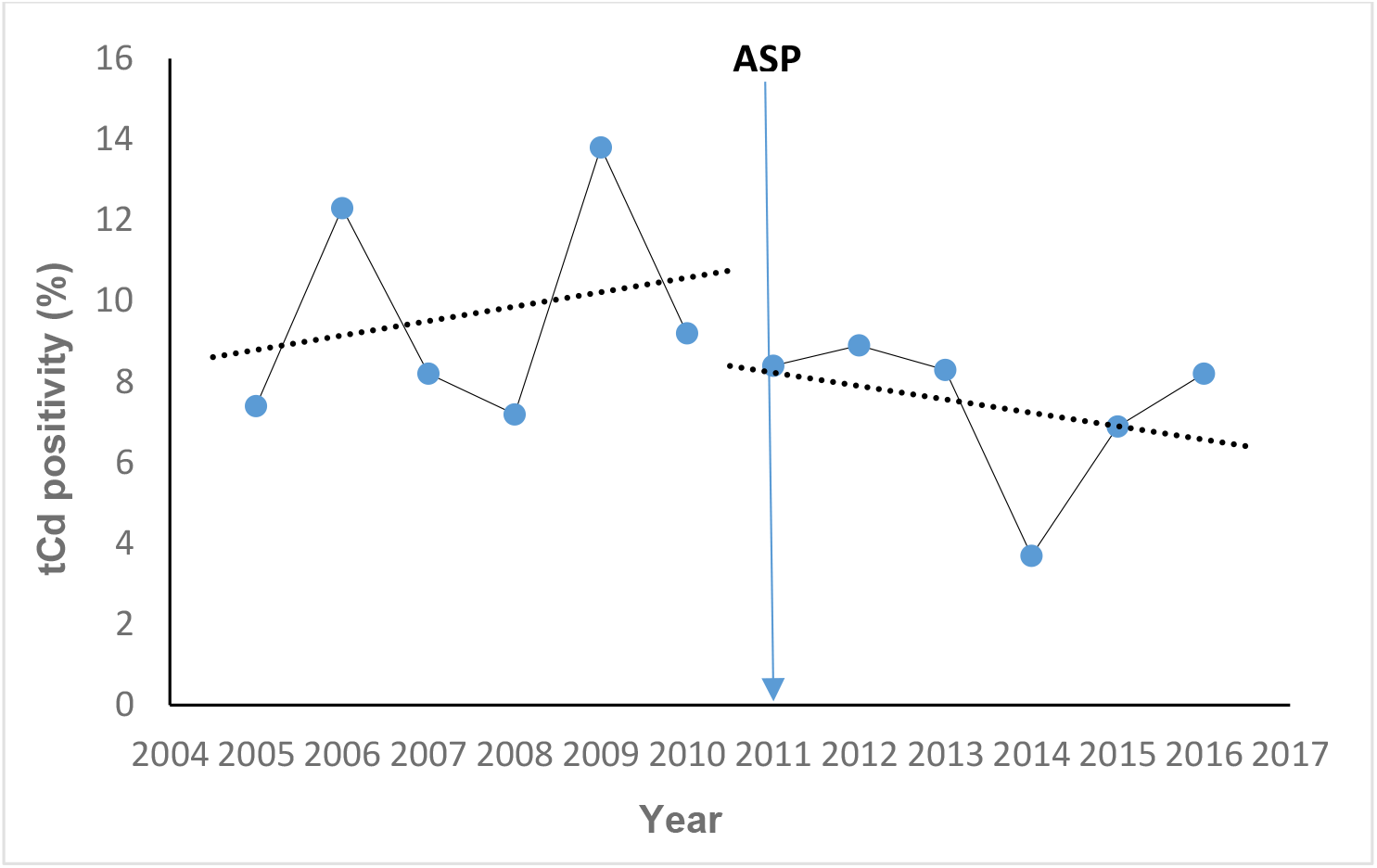
Secular trend in tCd positivity pre and post ASP

The decline in tCd positivity was observed in all three age groups, but this was statistically significant (p<0.05) only among paediatric patients (table 2 and figure 2).

**Table 2.** Age group-specific tCd positivity pre and post ASP

| Age group | number tested pre ASP | number positive pre ASP | Positivity (%) pre ASP | number tested post ASP | number positive post ASP | Positivity (%) post ASP | *P*-value |
| --- | --- | --- | --- | --- | --- | --- | --- |
| Elderly | 548 | 64 | 11.7 | 775 | 79 | 10.2 | .419 |
| Adult | 613 | 48 | 7.8 | 821 | 48 | 5.8 | .084 |
| Paediatric | 157 | 17 | 10.8 | 172 | 4 | 2.3 | .002 |
| Total | 1318 | 129 | 9.8 | 1768 | 131 | 7.4 | .022 |

Pre and Post ASP decline was significant ( $P < .05$ ) among paediatric patients

**Figure 2.**
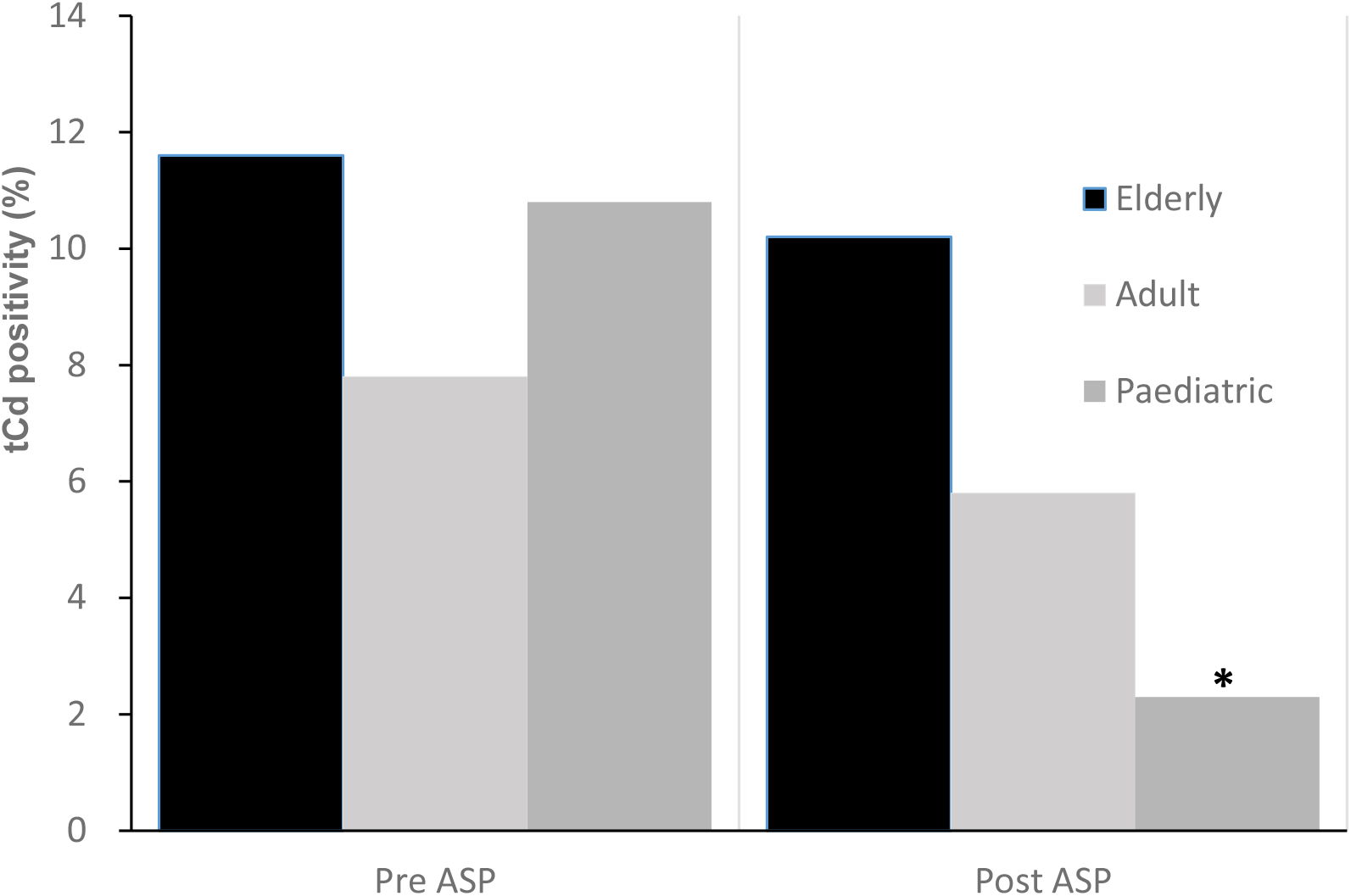
Age group-specific tCd positivity pre and post ASP. *Significant decline (p<0.05)

There were no significant differences in tCd positivity by gender, with females accounting for 139/260 (53%) and males 121/260 (47%) of the tCd positive patients identified.

## DISCUSSION

Unlike North America and Europe from where the majority of studies on CDI epidemiology have emerged, Saudi Arabia has a predominantly young population (<65 years), with the elderly (≥65 years) accounting for less than 5% of the population (United Nations, World Population Prospects, 2019). Therefore the predominance of the younger age groups among patients tested for C.diffiicle in the current study is consistent with this demographic distribution. The age-group distribution of tCd positive patients observed in the current study is also consistent with North American and European studies (McDonald *et al* 2006, Khanna *et al* 2012, Pechal *et al* 2016) that identify the elderly as being at especially high risk for CDI.

Although the post-ASP decline in tCd positivity was observed in all three age groups, this was statistically significant only among paediatric patients. The reason for this outcome is uncertain, but may be related to the excess burden among older patients of other CDI risk factors, including clinical co-morbidities and exposure to proton pump inhibitors and histamine 2 (h2) antagonists (Kwok *et al* 2012, Rotman and Bishop 2013, Seto *et al* 2014) The outcome may also reflect a higher level of compliance with antimicrobial stewardship recommendations among paediatricians, although there is no evidence in the current study to support this.

In a systematic review, Feazel *et al* (2014) observed that studies reporting the most impact of antimicrobial stewardship on CDI were those conducted in elderly settings, and pre-authorization of antibiotics and formulary restriction a part of the program. Therefore, a plausible explanation for the non-significant impact of the ASP on tCd positivity among the elderly in the current study is that this high-risk group was not specifically targeted for intervention. Furthermore, the ASP consisted primarily of persuasive interventions, and did not involve restriction of antibiotics. This meant that prescribers, largely, maintained autonomy and were not obliged to comply with stewardship recommendations.

The impact of the ASP on tCd positivity among elderly and adult patients may also have been dampened by the outbreak of MERS coV that occurred at the study setting in 2013, during the post-ASP period. MERS coV disproportionately affected these age groups (Al-Tawfiq et al 2014), and was associated with an increase in empiric and prophylactic antibiotic use (Momattin et al 2018). This may have contributed to the reversal in 2014 of the downward trend in tCd observed in the post-ASP period (figure 2)

Despite these drawbacks, the ASP was associated with a significant overall decline in tCd positivity. Perhaps, support of the program by an infectious disease (ID) physician played a vital role in influencing antibiotic prescribing practices, and contributed to the observed effect. This assertion is consistent with other CDI studies (Jump *et al* 2012, Tedeschi *et al* 2017, Ostrowsky et al 2018) highlighting the impact of ID physician support in antimicrobial stewardship, either alone, or in combination with other interventions..

The retrospective, before-and-after intervention study design limits the extent to which the observed outcome can be attributed solely to the ASP. The influence of other related interventions implemented before the ASP, that were on going over the study period cannot be discounted. These include the hand hygiene program, and the promotion of infection prevention and environmental services best practices for CDI. It is noteworthy however, that these interventions were in place prior to the ASP, and the rising trend in tCd period positivity was reversed only after the implementation of the program in 2011.

Life expectancy has increased in Saudi Arabia, and a more than 3-fold increase in the proportion of the elderly is projected over the next two decades (United Nations, World Population Prospects 2019). With this demographic shift, the prevalence of CDI is likely to increase. To mitigate this, there is a need for concerted efforts to implement antimicrobial stewardship in health care settings, and regulate population access to antibiotics. Periodic measurements of in-patient tCd positivity may offer a useful means of monitoring the impact of hospital antimicrobial stewardship programs, especially in those settings lacking the resources to accurately measure the rate of HA-CDI. The accuracy of this metric is however reliant on consistent best practices among clinicians and laboratory personnel. These include the avoidance of arbitrary repeated testing of a patient for cure, and testing only unformed or diarrhoea stool specimens. This will improve efficiency, reduce bias, and enhance the predictive value of a positive tCd test. Further studies are required within and beyond Saudi Arabia to examine the utility of this metric.

## Data Availability

Data referred to in this manuscript are held on the information management system of the Johns Hopkins Aramco healthcare Microbiology Laboratory, Dhahran, Eastern Province. Kingdom of Saudi Arabia

https://www.jhah.com/en/care-services/laboratory

